# Impact of airline travel network on the global importation risk of monkeypox, 2022

**DOI:** 10.1101/2022.09.17.22280060

**Authors:** Ryo Kinoshita, Miho Sassa, Shogo Otake, Fumi Yoshimatsu, Shoi Shi, Ryo Ueno, Motoi Suzuki, Daisuke Yoneoka

**Author notes:** **Correspondence** Daisuke Yoneoka, 1-23-1 Toyama, Shinjuku-Ku, Tokyo 162-0052 Japan.

## Abstract

**Background:** As of 4 September 2020, a total of 53,996 monkeypox cases were confirmed globally. Currently, most monkeypox cases are concentrated in Europe and the Americas, while many countries outside these regions are also continuously observing imported cases. We aimed to estimate the potential global risk of monkeypox importation and consider hypothetical scenarios of travel restrictions by varying passenger volumes via airline travel network.

**Method:** Passenger volume data for the airline network, and the time of first confirmed monkeypox case for a total of 1680 airports in 176 countries (and territories) were extracted from publicly available data sources. A survival analysis technique in which the hazard function was a function of effective distance was utilized to estimate the importation risk. Scenarios which selectively reduced the passenger volume from/to countries with detected monkeypox cases and increased/decreased the global passenger volume to the level of 2019 (high volume) or 2021 (low volume) regardless of monkeypox detection were considered for travel restrictions.

**Results:** The arrival time ranged from 9 to 48 days since the first case was identified in the UK on 6 May 2022. The estimated risk of importation showed that regardless of the geographic region, most locations will have an intensified importation risk by 31 December 2022. Travel restrictions had a minor impact on the global airline importation risk against monkeypox.

**Conclusions:** Instead of preventing the importation of monkeypox cases via airline networks, high risk countries should enhance local capacities for the identification of monkeypox and prepare to carry out contact tracing and isolation.

## Introduction

As of 4 September 2022, a total of 52,996 laboratory-confirmed cases of monkeypox has been identified globally [1]. The World Health Organization (WHO) declared the Public Health Emergency of International Concern (PHEIC) on 23 July [2]. Monkeypox virus (MPXV) belongs to the Orthopoxvirus genus and has been first detected as a human pathogen in the Zaire (currently known as the Democratic Republic of the Congo, DRC) in 1970. Recently detected endemic regions include; Cameroon, Central African Republic, DRC, Nigeria, Republic of the Congo [3–5]. Transmission to human occur through close contact with an infected animal or person or a contaminated material. Animal hosts include a variety of rodents and non-human primates, yet the exact reservoir of monkeypox is yet to be determined [6,7]. To present, monkeypox has not been considered to be contagious prior to symptom onset, while several case reports identifying asymptomatic infection raise concerns for the feasibility of controlling the multi-country outbreak [8,9]. Moreover, since the mean incubation period is estimated at 8.5 days and can be up to around 21 days, incidental importation can easily occur [7,10,11].

The risk of monkeypox has been continuously debated after the eradication of smallpox [5,12]. Despite knowledge of smallpox vaccination being effective against monkeypox, local and global smallpox eradication by 1980 lead to cessation of routine vaccination, and especially cohorts born post smallpox eradication is presumed to have no immunity against monkeypox [10,12]. Therefore, increased susceptibility to monkeypox infection especially among younger generations which were born after smallpox eradication may pose a higher risk to infection. Recently, several smallpox vaccines have become registered as a vaccine to protect against monkeypox [13]. The first documented outbreak of monkeypox outside of Africa was in the United States of America in 2003 with exposure from prairie dogs, and only several detected events of importations have occurred globally due to travelers arriving from endemic areas and none of them lead to sustained local transmission [6,14]. The only human-to-human transmission reported outside of Africa was in the United Kingdom (UK) in 2018 [15].

Considering the global travel network, large European cities has had a high risk of importing Monkeypox [16]. The current multi-country outbreak in 2022 was first identified in the UK, and sustained local transmission has been mainly observed in Europe and the Americas [3]. Moreover, the disease has been continuously identified in a majority of global locations. The first case was detected on 6 May 2022 in the UK due to importation from Nigeria [3,17]. While the index case still remain unclear, intra- and international spread of the disease has been observed with evidence of sustained human to human transmission [17–20]. As of 4 September, countries which have reported a high cumlative number of cases (>3000) globally were the United States of America (n = 19,351), Spain (n = 6645), Brazil (n = 5197), France (n = 4646), Germany (n = 3493) and the United Kingdom (n = 3413) [1]. Outside of African region, 8 deaths have been confirmed to present [1]. A unique characteristic of the current spread of monkeypox cases is that most cases are concentrated among young men, in the population of men who have sex with men (MSM) [1,15,18,21]. Typically monkeypox is not considered to be a sexually transmitted infection, while it can be transmitted easily during sexual and intimate contact [22]. While global international travel volume has been largely reduced from 2020-2021, in the present year, the International Air Transport Association (IATA) expects air passenger volume to be 69% compared to 2019 (pre-COVID-19 pandemic) [23]. Due to resumptions of human movement via airline transportation, travelers could unintentionally cross the border with monkeypox infection.

Several mathematical modeling techniques have been developed responding to the rapid dissemination of emerging infectious diseases fueled by airline travel network [24–31]. Practically travel restrictions have been put in place in several boarders during the COVID-19 pandemic, specifically when it was discovered in Wuhan, China and also when a new variant of concern has been identified (i.e. the Omicron variant in South Africa) [32–34]. Quantifications of the impact of travel restriction against the risk of importation has been simulated using airline transportation network arriving from Wuhan, China [35]. Several studies have also quantified the delayed epidemic progression given the stringent travel restriction in Wuhan, China [36,37].

The present study modified the model previously applied against COVID-19 for the quantification of risk of importation [35]. Our study aimed to quantify the global risk of importation using airline transportation data. Using a hazard-based model and the concept of effective distance based on travel network data, we explored patterns of domestic and international population movement. Our findings on travel patterns could contribute to inform public health interventions, especially for understanding the risk of observing an emerging disease across the border.

## Methods

### Dataset and global airline network

The dates of first onset of monkeypox case for each location (including country, city and/or name of hospital) were extracted from open-access database developed in response to the multi-country outbreak [38,39] at 5 September 2022. The data included 175 countries or territories. Two authors, RK and DY, independently checked the validity of data against official announcements from WHO and each governmental report. The extracted case information was then matched with the airport information based on the nearest neighborhood approach. To construct the airline transportation network, we used the ADS-B exchange data [40]. The dataset provides a graph of global travel information consisting of 1,724 nodes (corresponds to each airport) and 21,704 edges with edge-weights (corresponds to direct flights between two airports with its passenger volume (PV). The PV on a certain flight was estimated as the reported (maximum) number of seats of the airplane. Then, the PV was multiplied by 0.93 for domestic travel and 0.69 for international travel [23].

### Effective distance

To model the impact of airline network on monkeypox transmission, an idea of *effective distance*, which was introduced by Brockmann and Helbing [41] and frequently used in previous studies for forecasting the global spread of emerging infectious disease such as SARS, influenza H1N1-2009, MERS and COVID-19 [29,35], was estimated from the airline network. The effective distance is defined from the minimum distance on the adjacency matrix of the network, incorporating the PV-weighted path length and the degree of each node.

The effective distance, *d*_*ij*_, between London Heathrow airport (ICAO code: EGLL) and the *i*th airport in the *j*th country is defined as the minimum length among all possible effective paths. The effective paths from Heathrow airport to the *i*th airport with a sequence of *l* transit airports {*a*_*Heathrow*_ *a*_1_, …, *a*_,*l*−1_, *a*_*i*_}is given by

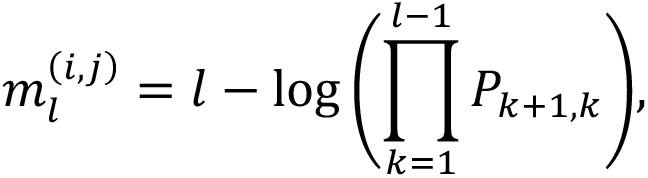

where *P*_*l,m*_ denotes the transition probability matrix from the *l*th to the *m*th airport. Each element in *P*_*l,m*_ is estimated by 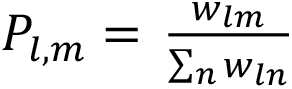, where *w*_*lm*_ is the PV that moved from the *l*th to the *m*th airport. Lastly, *d*_*ij*_ is defined as the minimum effective paths, which is given by

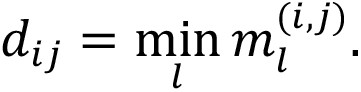

As we will discuss in the next section, since the network structure and its associated effective distance changed after the travel restrictions due to the change in the PV, the effective distance *d*_*ij*_ is dynamically changed in the assumed three scenarios described below.

### Modeling with effective distance: hazard-based approach

We modeled the risk of importing monkeypox by estimating the survival probability in each country. Let *T* be a random variable indicating the survival time in each airport from the first case in the UK (6 May 2022) to importation at the airport. Also define the survival probability at time of *t* as *F*(*t*) = *P*(*T* > *t*) with the probability density function (pdf) of *f*(*t*) = −*dF*(*t*)/*dt. t* indicates day, and *t* = 0 is 6 May 2022. The associated hazard function at time of *t* for importation of monkeypox from the UK for the *i*th airport in the *j*th country is modeled in the form of Weibull regression model, which is given by

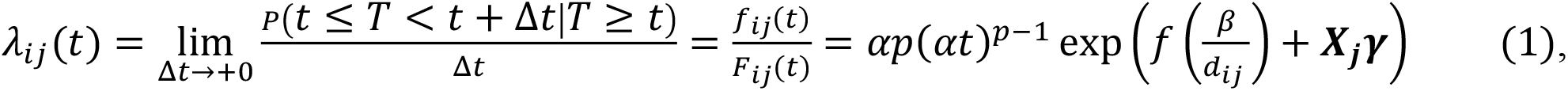

where *p* > 0, *α* > 0, ***γ*** is a vector of regression parameters, *β* is also a regression parameter of interest to measure the impact of the effective distance, *f*() is a penalized smoothing spline function with the degree of freedom of 4, and ***X***_***j***_ is a covariate vector. This formulation makes the hazard function and the estimated median time of importation be proportional to the effective distance *d*_*ij*_, which is consistent with Shi *et al*. (2021) and Otsuki and Nishiura (2016) [29,35]. The covariates ***X***_***j***_ include: income per capita at 2022 [42], the proportion of working age at 2020 [43], the proportion of sexual minorities concealing their sexual orientation [44], the proportion of MSM population [45,46], Socio Demographic Index at 2019 [42] and WHO region (i.e., African Region (AFR), Eastern Mediterranean Region (EMR), European Region (EUR), Region of the Americas (AMR),

South-East Asian Region (SEAR), Western Pacific Region (WPR)). Naturally, Equation (1) can be rewritten as the pdf of survival time as

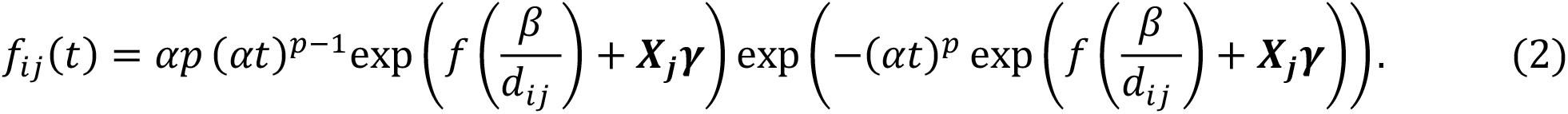

The parameters were estimated by the maximum likelihood approach. Then, the future hazard function at 31 December 2022, was predicted by extrapolating the time variable *t*. In order to capture the importation risk from the UK, the following countries currently designated as endemic of monkeypox is not included in the parameter estimation: Benin, Cameroon, the Central African Republic, the Democratic Republic of the Congo, Gabon, Ghana, Ivory Coast, Liberia, Nigeria, the Republic of the Congo, Sierra Leone, and South Sudan [3]. Further, to check the goodness-of-fitting in the model, we calculated the concordance index, which measures the agreement between an observed response and the predictor.

Lastly, we conducted the sensitivity analysis by weakening the assumption in the effective distance: i.e., since the effective distance strongly depends on the assumption that monkeypox cases has been spreading from the UK, we checked how our result would be changed when weakening this assumption (i.e., not specifying the starting point of the virus spread). More precisely, we used a “closeness centrality index” on the airline network, which measures how attractive a certain airport is in the PV sense, instead of the effective distance in the model. The closeness centrality for the *i*th airport is defined as

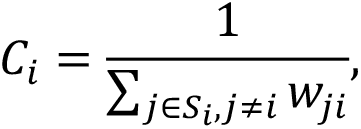

where *w*_*ji*_ is the PV that moved from the *j*th to the *i*th airport and *S*_*i*_ is the set of airports that are connected to the *i*th airport. Then, instead of *d*_*ij*_ in Equation (2), *C*_*i*_ is used. *C*_*i*_ does not assume the starting point of the virus spread, and in that sense we do not make the assumption that the infection is spreading from the UK in this sensitivity model.

### Hypothetical scenarios to estimate relative risk reduction due to travel restriction

To estimate the impact of travel restrictions on the importation risk, we calculated the (observed) cumulative risk at time of *t*, defined by

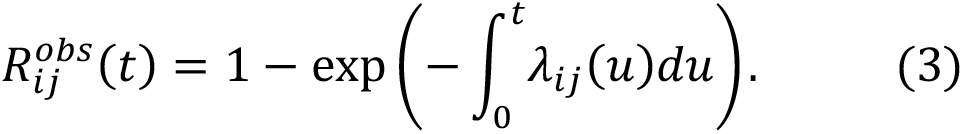

Then we compared 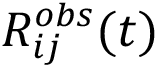 with the cumulative risk of following hypothetical scenarios: (H1) Reduce the current PV from/to infected countries by 50%, (H2) Increase the current PV to “pre-COVID-19 level in 2019” (cf. the current PV was assumed 93% for domestic and 69% for international travel, respectively, compared with the pre-COVID-19 level) [23], and further reduce the PV from/to infected countries by 90%, and (H3) Reduce the current PV to “the level at the most severe travel restrictions in 2021” (i.e., the PV is assumed 61% for domestic and 27% for international travel, respectively, compared with compared with the pre-COVID-19 level), and further reduce the PV from/to infected countries by 50% [23]. Once the regression parameters in Equation (1) was estimated, the cumulative risks based on these scenarios can be calculated by plugging the scenario-specific *d*_*ij*_ into Equation (3): i.e., in a similar way with the calculation of 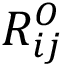, the cumulative risks in (H1)-(H3) are given by

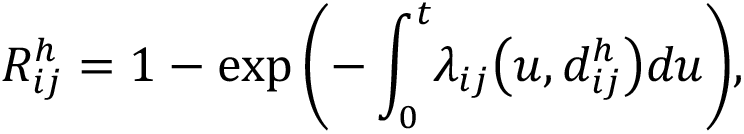

where *h* =1, 2, 3 indicates scenarios (H1)-(H3), respectively, and 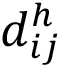 is the effective distance based on each scenario assumption. Lastly, we estimated the relative risk change as follow:

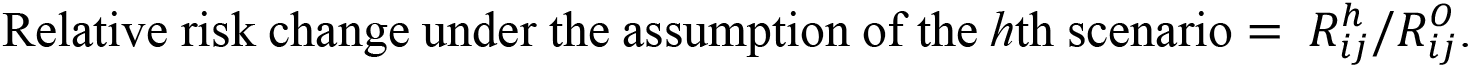

To measures the proportion of expected risk reduction between observed and the hypothetical scenarios the relative risk change was calculated at *t* = 239 (31 December 2022).

## Results

Figure 1(A) shows the entire flight network with the associated PV. PV tends to be higher in intercontinental flights compared to intracontinental flights. Figure 1(B) indicates PV arriving/departing from the UK and Heathrow Airport. A total of 176 countries (and territories), including the UK, and 1680 airports were included in this analysis. The arrival time ranged from 9 to 48 days since the first case was identified in the UK on 6 May 2022.

**Figure 1.**
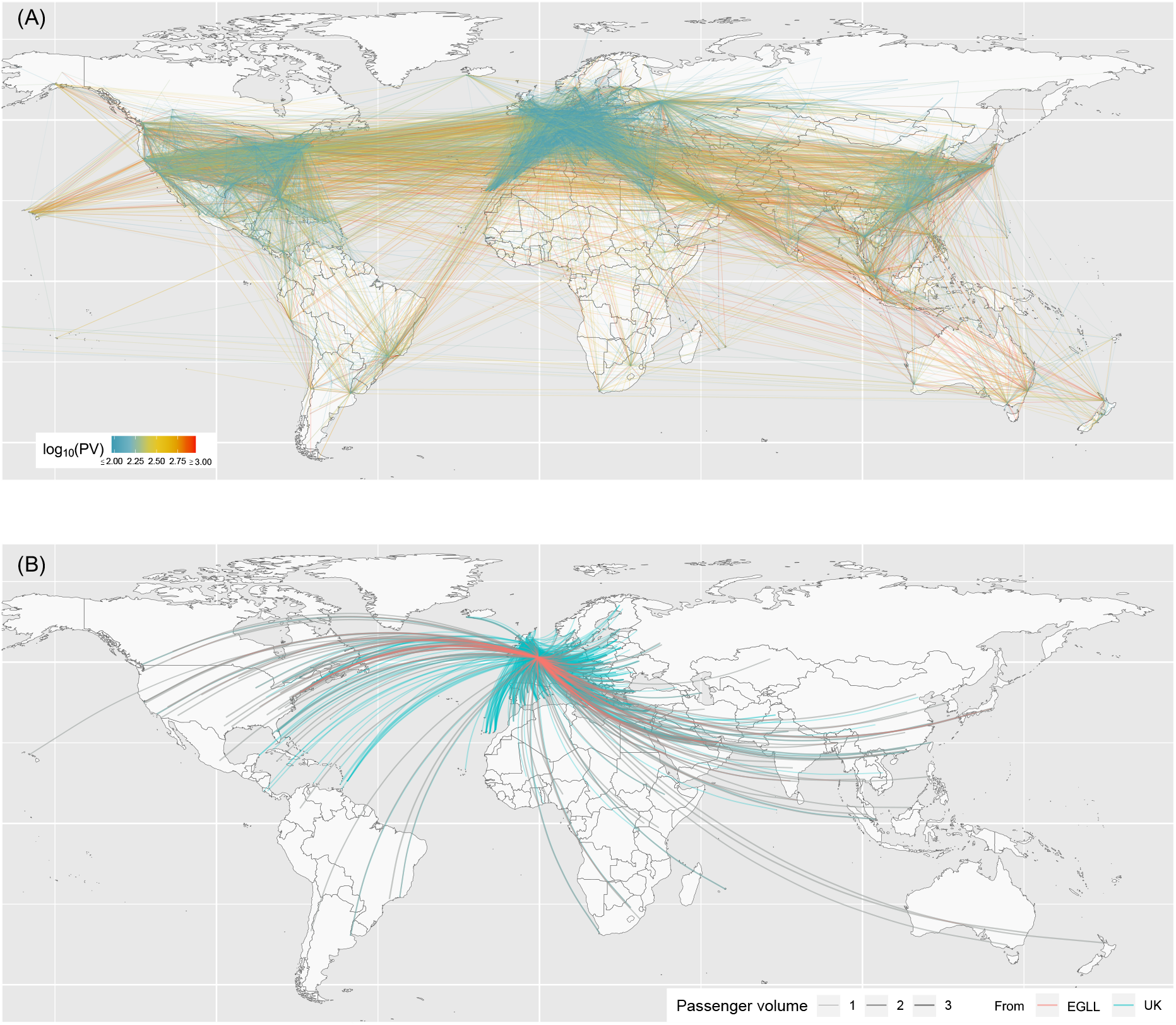
(A) Entire flight network before travel restrictions (as of 1 December 2019). Color indicates the passenger volume (PV) in log scale. (B) Flight network from the UK and Heathrow Airport (as of 1 December 2019)

Figures 2(A) and 2(B) show the estimated and predicted cumulative risk 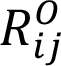 at 5 September and 31 December 2022, respectively. The increased risk over time was similar between WHO regions; the median risk ratios (defined as 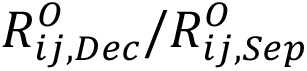) and interquartile range (IQR) in AFR, EMR, EUR, AMR, SEAR, and WPR were 1.56 (IQR = 0.0305), 1.57 (IQR = 0.0593), 1.52 (IQR = 0.0860), 1.50 (IQR = 0.0997), 1.58 (IQR = 0.0196), and 1.58 (IQR = 0.0157), respectively. The estimated concordance index was 0.816, which implicates the model has good fitting ability. To check the robustness of our result, we note that similar results were obtained even in the sensitivity analysis (Supplemental Figure).

**Figure 2:**
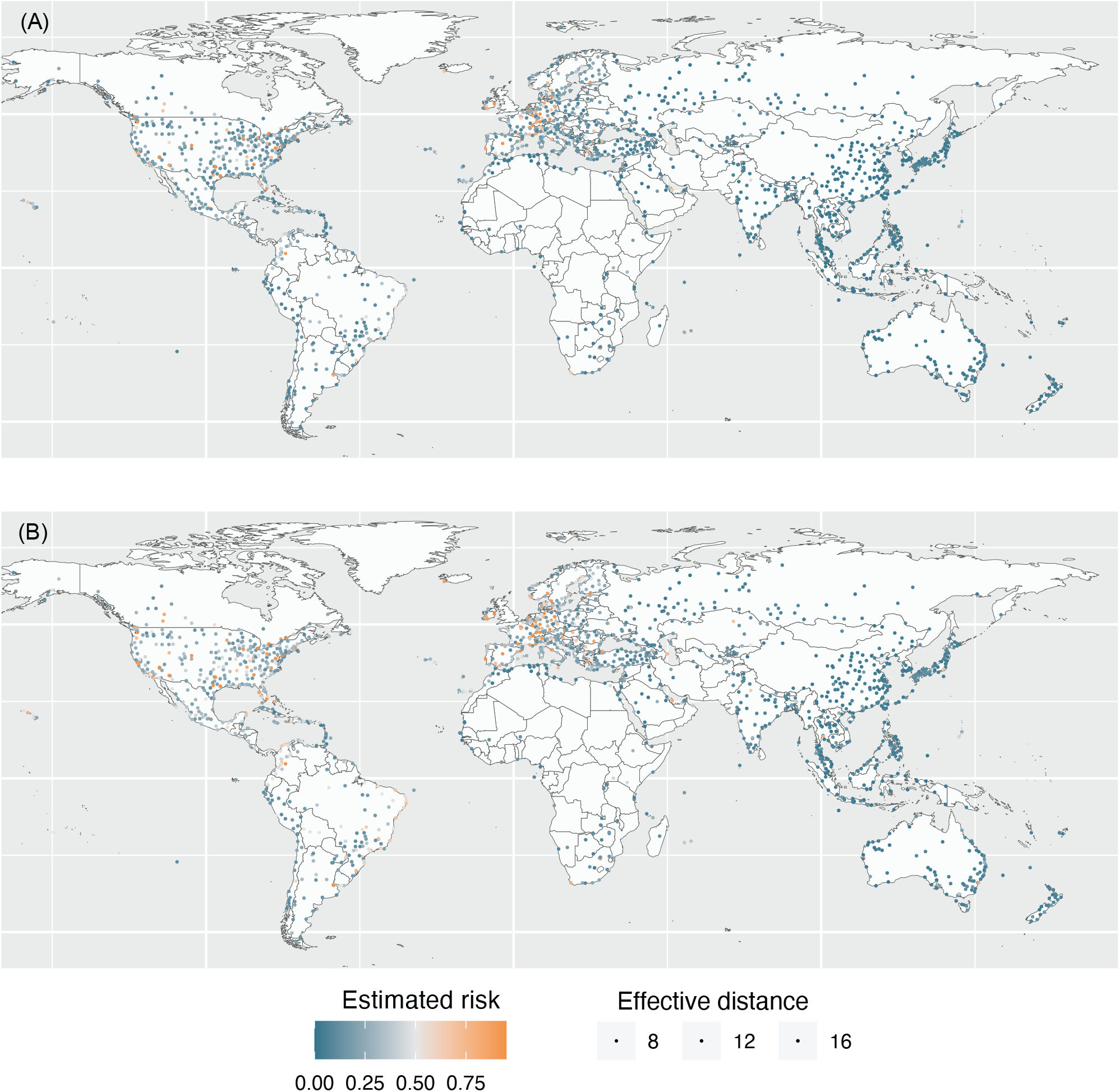
(A) Estimated risk of importation of monkeypox as of 5 September 2022. Color indicates the estimated risk and the size of circles indicates the number of inbound passenger volume of each airport. (B) Predicted risk of importation of monkeypox as of 31 December 2022.

Figures 3(A), 3(B) and 3(C) show the estimated relative risk change under the assumption of (H1)-(H3). The mean (Standard deviation (SD), max and min) for the relative risk change were 0.968 (SD=0.075, max = 1.195, min = 0.766), 0.953 (SD=0.240, max = 1.982, min = 0.433) and 0.963 (SD=0.113, max = 1.291, min = 0.685) for (H1)-(H3), respectively. Regarding the geographical distribution of the risk change, in (H1), there was characteristic risk decrease, especially in Europe, Scandinavia, Southeast Asia, and Australasia, while there was risk increase in Central America and the Caribbean countries. In (H2), where the overall PV is assumed to return to “pre-COVID-19 level”, importation risk increase is observed at similar locations as observed in (H1), while the increase was stronger due to increased travel volume. Surprisingly, areas observing risk decrease was also similar to (H1), and the decrease was stronger even despite increased overall PV. In (H3), risk increase is prevented compared to (H2) due to heavy reduction in the PV assuming 2021 level. However, the strong risk decreases as observed in (H2) is not observed even with significant decrease in overall PV. Interestingly, in all intervention scenarios Central America had an increased risk. Detailed values by countries are provided in the Supplementary Table.

**Figure 3:**
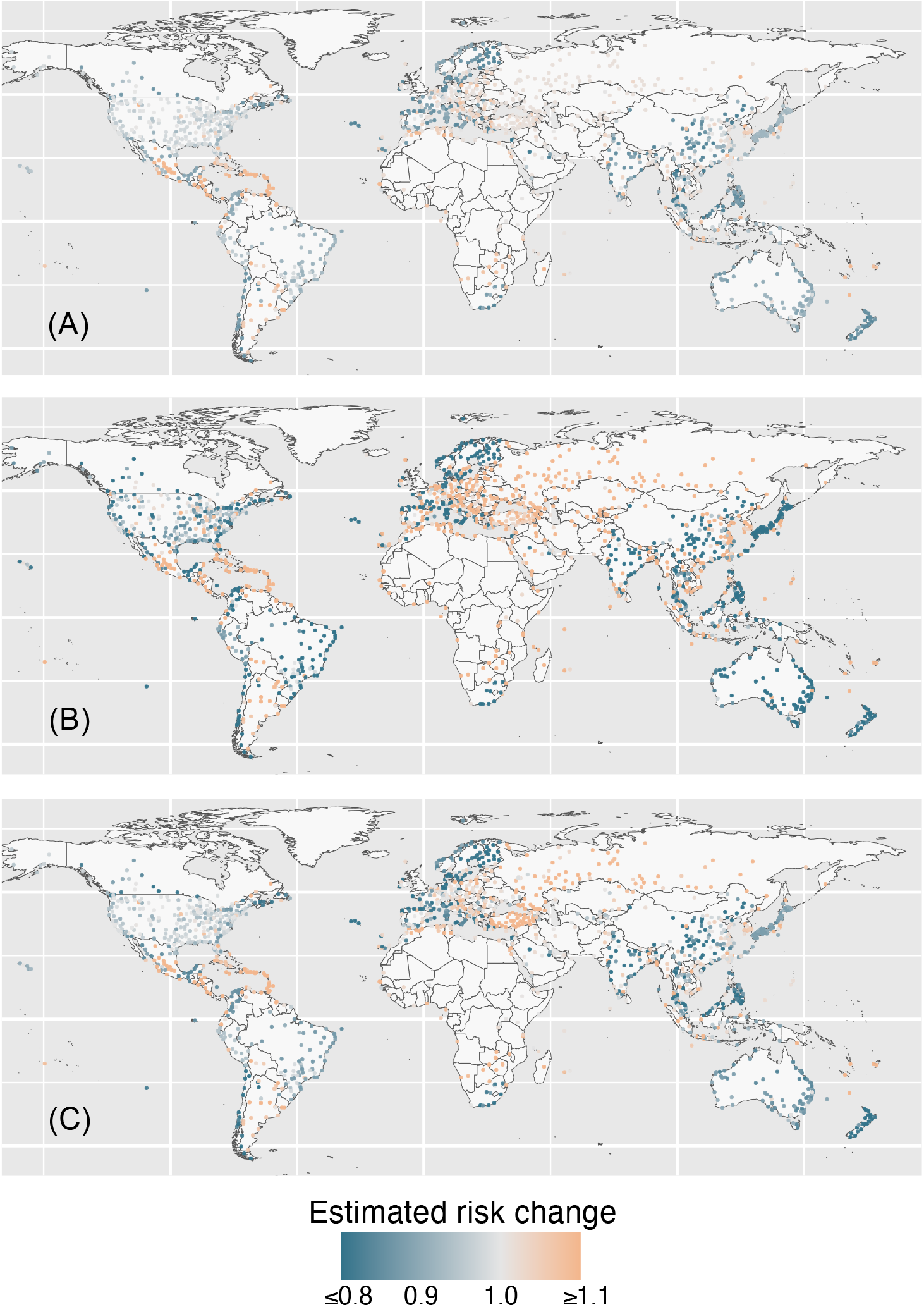
(A) Predicted relative risk change under the assumption of H1 as of 31 December 2022. Color indicates the estimated relative risk change. (B) Predicted relative risk change under the assumption of H2 as of 31 December 2022. (C) Predicted relative risk change under the assumption of H3 as of 31 December 2022.

## Discussion

Employing a hazard based model and utilizing the idea of effective distance, the present study estimated the importation risk against monkeypox. Assuming that the current flight volume would be maintained, the risk of importation by 31 December 2022 is expected to be substantial in multiple locations, including areas which yet have experienced sustained local transmission (Figure 2). Strong risks are seen in locations with large PV with the presence of a closely connected flights irrespectively to the actual distance from London, UK. Our result highlights the importance to enhance surveillance against monkeypox in nations with a high risk of importing monkeypox.

To confirm a hypothetical scenario where travel restrictions are imposed, we reduced PV from/to countries already detected to have monkeypox importation. The reduction of PV by 50% may be considered as a situation when travel recommendations were made not to travel in these locations unless necessary (H1 and H3). The reduction of PV by 90% may be considered as a situation when strict travel restrictions were implemented to/from countries identifying monkeypox (H2). The relative risk reduction given travel restriction only had a minor effect to the risk of importation (Figure 3A). To verify the sensitivity to different PV on the relative risk reduction, we changed the passenger volume from/to identified location, and the degree of reduction in overall PV considering the travel volume in 2019 (pre-COVID-19 level; high PV) and 2021 (least travel volume during the COVID-19 pandemic; low PV). While minor changes in the risk is observed, varying the global airline PV and travel restrictions from/to identified locations did not strongly contribute to modify the risk of importation (Figure 3BC), suggesting that the degree of PV has a nonlinear effect on the risk reduction, and the optimal size of volume reduction may depend on the connectivity between each airport network as also discussed previously [35]. Therefore, our hypothetical scenarios examined that practical implementation of travel restrictions or recommendations to reduce PV in order to minimize the risk of importing monkeypox cases may not be an efficient strategy. Rather, intensified contact tracing and isolation would be more important once a single case is identified in a new location.

Several limitations must be discussed. First, this study estimated the probability of importation using only airline network data, while sea and ground (automobiles and railway) network are also drivers to human mobility. Second, our analysis does not take into account of the change in the airline travel network structure due to humanitarian crisis occurring between Ukraine and Russia. Third, we defined that the spread of monkeypox originated from London, UK and estimated the global importation risk. However, similar results were obtained in our sensitivity analysis relaxing this assumption. Fourth, our analysis focused on the risk of importation, and therefore the risk of local transmission given importation is not quantified. Despite these limitations, our projection exercise highlights the propagating global risk of importation of monkeypox cases using airline transportation network.

In conclusion, travel restrictions can impose strong economic and social impact, and thus careful evidence-based decision process is necessary [33]. While our simple model may not fully capture the complex dynamics of global disease transmission, our simulation showed that in the case of monkeypox, airline travel restrictions may not be the practical intervention to prevent importation in most areas. Instead of preventing the importation of monkeypox cases via airline networks, countries especially considered to have a high risk of importation should enhance local capacities for the identification of monkeypox and prepare to carry out contact tracing and isolation.

## Data Availability

The data underlying this article and R programs will be shared on reasonable request to the corresponding author.

## Abbreviations

PV: passenger volume

## Footnotes

### Author Contributions

RK and DY led the study. All authors took responsibility for the integrity of the data and the accuracy of the data analysis. All the authors made critical revisions to the manuscript for important intellectual content and gave final approval of the manuscript. The opinions, results, and conclusions reported in this paper are those of the authors and are independent from the funding bodies.

### Conflict of interest

The authors have declared no conflicts of interest.

## Supplementary materials

**Supplementary Figure:**
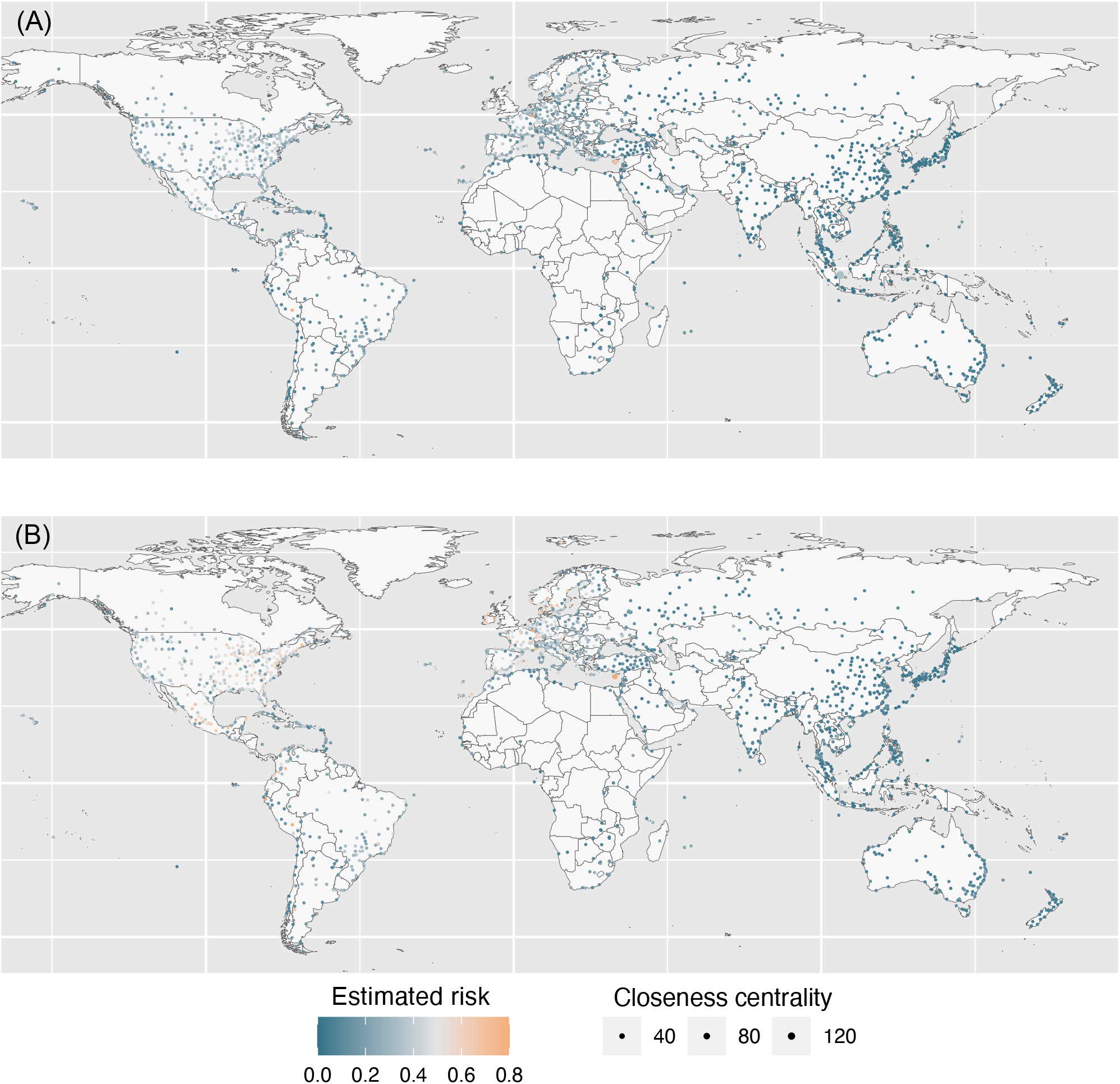
(A) Estimated risk of importation of monkeypox as of 5 September 2022 using network closeness centrality. Color indicates the estimated risk and the size of circles indicates the estimated closeness centrality. (B) Predicted risk of importation of monkeypox as of 31 December 2022 using network closeness centrality.

Supplementary Table: Detailed estimated values of Figures 2 and 3. Shown in parenthesis is the standard deviation (SD).

## Notes

**Funding:** This work was supported by JST, PRESTO Grant Number JPMJPR21RC, Japan, and the Japan Society for the Promotion of Science (JSPS) KAKENHI (21K17307).

### Competing Interest Statement

The authors have declared no competing interest.

### Funding Statement

This work was supported by JST, PRESTO Grant Number JPMJPR21RC, Japan, and the Japan Society for the Promotion of Science (JSPS) KAKENHI (21K17307).

